# Biological age acceleration and the dynamic progression of cardiovascular-kidney-metabolic diseases to multimorbidity, dementia and mortality: A prospective cohort study

**DOI:** 10.64898/2026.04.07.26350289

**Authors:** Guojing Yuan, Ping Zeng

**Author notes:** Correspondence to: Ping Zeng, Department of Biostatistics, Jiangsu Engineering Research Center of Biological Data Mining and Healthcare Transformation, and Jiangsu Key Laboratory of Geriatric Precision Medicine and Aging Intervention, Xuzhou Medical University, Xuzhou, Jiangsu 221004, China.

## Abstract

**Background:** The role of biological age acceleration (BioAgeAccel) in the dynamic progression from single cardiovascular-kidney-metabolic disease (CKMD) to multimorbidity, and subsequently to dementia and mortality remains elusive.

**Methods:** We conducted a longitudinal study with data of 433,911 UK Biobank participants. Cardiovascular-kidney-metabolic multimorbidity (CKMM) was defined as the coexistence of two or more CKMDs, including cardiovascular disease (CVD), stroke, type 2 diabetes (T2D), and chronic kidney disease. Biological aging was measured via PhenoAge and KDM-BA. Multistate models examined the association between BioAgeAccel and disease transitions, ranging from healthy to the first occurrence of CKMD (FCKMD), then progression to CKMM, dementia, and mortality. Restricted mean survival time estimated the disease transition time or life expectancy between states.

**Results:** BioAgeAccel was significantly associated with increased risks across all disease transitions. Specifically, during CKMM progression, the hazard ratios (HRs) of the transition from healthy to FCKMD were 1.24 [95%CI 1.23-1.25] for PhenoAgeAccel and 1.16 [1.15-1.17] for KDM-BA-Accel. For subsequent transition to CKMM, the HRs were 1.20 [1.18-1.22] and 1.19 [1.17-1.21], respectively. In dementia-related transitions, PhenoAgeAccel showed the higher risk for CKMM to dementia (HR=1.13 [1.04-1.22]) than for the transition from healthy or from FCKMD to dementia. These associations were further moderated by age, physical activity, educational, and lifestyle factors. BioAgeAccel also accelerated disease progression and reduced life expectancy; for example, during CKMM progression, BioAgeAccel shortened the time between disease transitions by about 1.09 years from healthy to FCKMD, and an additional 1.75 years to CKMM. Regarding life expectancy, individuals with CKMM experienced an average reduction of about 1.36 years under PhenoAge, while those with dementia showed a decrease of about 0.77 years. Among individuals with CVD or T2D as the initial diagnosis, the impact of BioAgeAccel on progression to CKMM or dementia was stronger.

**Conclusions:** BioAgeAccel exerts significant promotive role in the onset of CKMD and their subsequent progression to CKMM, dementia, and mortality, helping identify high-risk individuals. Implementing biological age assessments and health lifestyle interventions in middle-aged populations serves as an effective strategy for alleviating the burden of CKMDs and dementia.

Graphical abstract

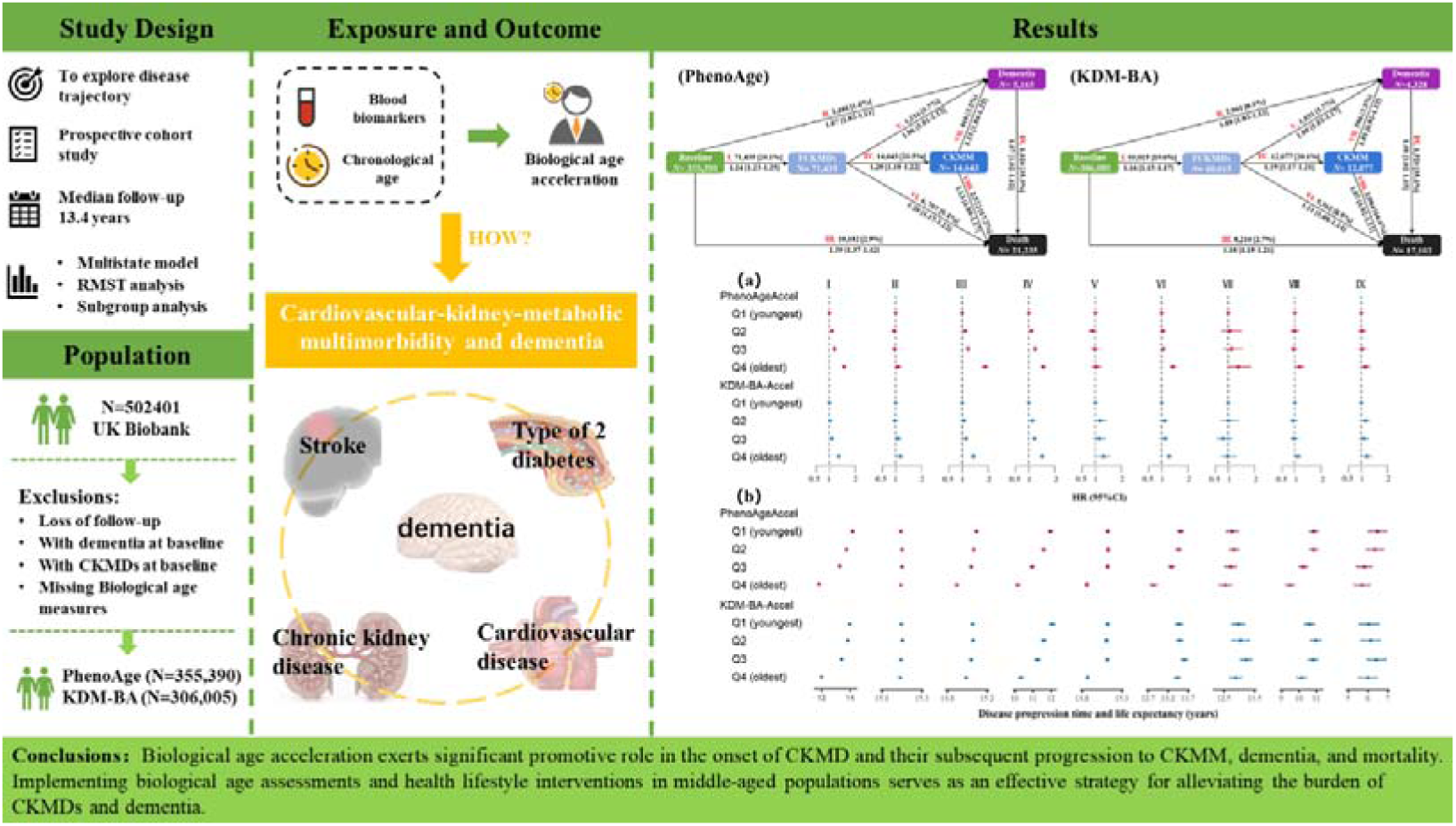

## Introduction

Dementia and cardiovascular-kidney-metabolic diseases (CKMDs) are two major global public health priorities that urgently require attention. With the accelerating trend of population aging, the prevalence of dementia is rising rapidly (Cook, et al., 2019). Currently, approximately 50 million people worldwide are living with dementia, with nearly 10 million new cases each year (World Health Organization., 2017), posing a serious threat to the health and life quality of the elderly population. Meanwhile, CKMDs, increasingly recognized as an integrated disease framework encompassing a cluster of interrelated chronic conditions including type 2 diabetes (T2D) mellitus, chronic kidney disease (CKD), cardiovascular disease (CVD) and stroke (Ndumele, et al., 2023), have become one of the leading causes of global mortality and disability, accounting for about one-third of deaths in the world (Ostrominski, et al., 2023; Zhang, et al., 2024). Moreover, extensive research has discovered that a variety of CKMDs are independent risk factors for dementia (Barbiellini Amidei, et al., 2021; Miwa, et al., 2014; Wolters, et al., 2018; Zheng, et al., 2024).

With substantial gains in life expectancy and advancements in chronic disease management, individuals initially diagnosed with first CKMDs frequently progress to cardiovascular-kidney-metabolic multimorbidity (CKMM) over time (Marassi and Fadini, 2023; Ostrominski, et al., 2023). For instance, results from the National Health and Nutrition Examination Survey indicates that approximately 49.0% of adults have at least one metabolic risk factor or moderate-to-high-risk CKD, with 14.6% eventually developing CKMM (Aggarwal, et al., 2024). Notably, multimorbidity exerts a synergistic effect on dementia risk; for example, compared to individuals with one single disease, those with at least two cardiometabolic conditions face a 68% higher risk of encountering dementia (Dove, et al., 2023). These findings underscore the significance of identifying and targeting commonly modifiable risk factors of these two types of diseases to delay the progression of CKMDs and mitigate the associated risk of dementia.

Importantly, CKMDs and dementia, to a great extent, share common pathophysiological mechanisms, such as chronic inflammation, oxidative stress, and vascular stiffening — processes that are largely driven by biological aging (Kennedy, et al., 2014; Wang and Bennett, 2012). In recent years, various novel biological aging biomarkers based on clinical parameters have emerged, offering a systematic reflection of aging across multiple organs or systems (Cesari, et al., 2017). Among these, algorithms such as PhenoAge (Liu, et al., 2018) and Klemera-Doubal Method Biological Age (KDM-BA) (Klemera and Doubal, 2006) that integrate multiple clinical and physiological indicators, have been validated for their high accuracy in predicting disease incidence and mortality risk (Belsky, et al., 2018; Li, et al., 2020). Recent studies found that accelerated biological aging was significantly associated with increased risk of dementia (Jiang, et al., 2025) as well as distinct CKMDs (Wang, et al., 2025; Zhao, et al., 2024; Zheng, et al., 2025). These findings suggest that biological aging serves as a key driving force in the dynamic progression of CKMDs and their transition toward dementia; however, its associations with the dynamics of CKMM development and their progression to dementia and mortality has yet to be investigated.

To address this research gap, by leveraging large-scale prospective cohort data from the UK Biobank (Sudlow, et al., 2015), this study seeks to examine the impact of biological aging on the onset of CKMDs and its transitions to CKMM, dementia, and mortality using multistate models (Putter, et al., 2007). To calculate biological aging, we employed the PhenoAge (Liu, et al., 2018) and KDM-BA algorithms (Klemera and Doubal, 2006). Methodologically, compared to traditional survival analysis methods, multistate models allow for simultaneous consideration of multiple intermediate states and transition pathways (e.g., single disease → multimorbidity → dementia), thereby offering a more realistic representation of disease evolution in populations with multimorbidity. This makes multistate models particularly valuable in the study of complex chronic conditions.

In summary, our study is the first to investigate, from a dynamic disease progression perspective, to assess the role of biological aging as a driving force in multistage disease trajectories. The findings are expected to provide theoretical insights for optimizing dementia prevention strategies and contribute to alleviating the global public health burden of CKMDs and dementia.

## Methods

### Study design and participants

This study was conducted with data from the UK Biobank (Sudlow, et al., 2015), a prospective community-based cohort that recruited over 500,000 residents aged 37-73 years across the United Kingdom between 2006 and 2010. Through standardized questionnaires, physical measurements, and biological sample collection, the cohort systematically collected comprehensive demographic characteristics, lifestyle factors, and health-related parameters.

From the original cohort, we first excluded individuals lost to follow-up (*N*=1,298) and those diagnosed with dementia at baseline (*N*=216), obtaining 500,887 dementia-free participants (Figure 1). Then, to assess the role of biological aging in the onset and progression of CKMD to CKMM, dementia, and mortality, we removed participants with CKMD at baseline (*N*=66,976). Based on the completeness of biomarker data, we further refined the analytic samples: for PhenoAge, we ruled out 78,521 participants with missing biomarker data, and the resulting sample size was 355,390; for KDM-BA, we excluded 127,906 participants, and ultimately kept 306,005 samples.

**Figure 1.**
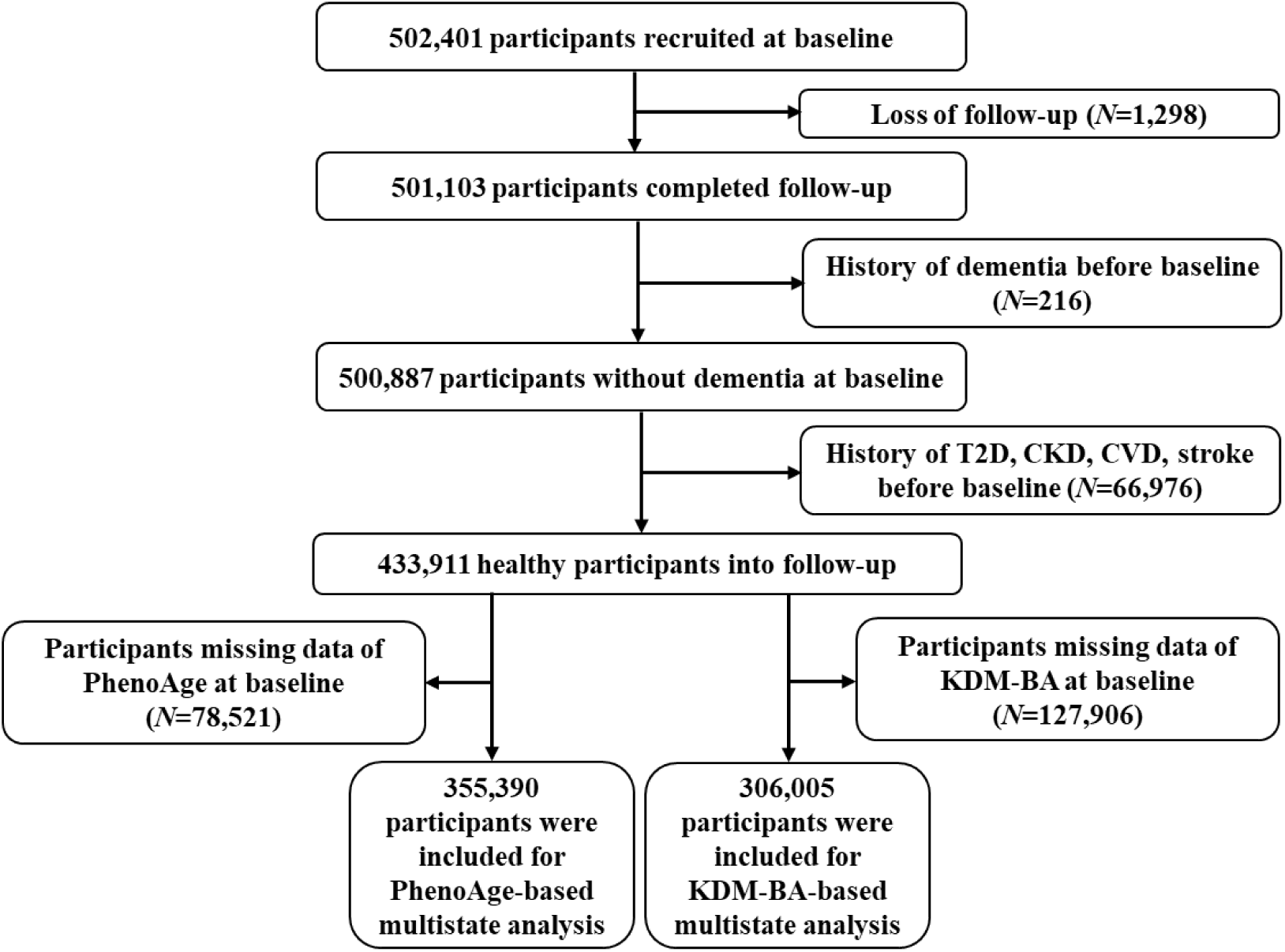
Flowchart of participants included in this study. T2D, type 2 diabetes; CKD, chronic kidney disease; CVD, cardiovascular disease.

### Assessment of CKMDs, dementia, and all-cause mortality

We primarily identified cases of CKMDs, dementia, and mortality through hospital inpatient data and death registration records, with self-reported data and primary care data used as supplementary sources. The follow-up period was calculated from the date of recruitment until the diagnosis of outcomes, date of death, or the censoring date of July 19, 2021, whichever occurred first.

We defined the study endpoint events based on ICD-10 codes, including CVD (I20-I25, I48, I50, I70, I73), stroke (I60-I69), T2D (E11, E14), CKD (N18), and dementia (F00-F03, G30). Based on previous studies (Jiang, et al., 2024; Ndumele, et al., 2023), we categorized CKMDs into four major groups: CVD (including coronary heart disease, atrial fibrillation, heart failure, peripheral vascular disease), stroke, T2D, and CKD. CKMM was defined as the coexistence of at least two of the aforementioned four disease categories.

### Assessment of biological aging and biological age acceleration

Biological aging was measured via two well-validated algorithms: the PhenoAge algorithm (Liu, et al., 2018) and the KDM-BA algorithm (Klemera and Doubal, 2006). PhenoAge was calculated through a linear combination of chronological age and nine blood biomarkers, including albumin, alkaline phosphatase, creatinine, C-reactive protein, glucose, mean corpuscular volume, red cell distribution width, white blood cell count, and lymphocyte percentage. KDM-BA was derived from forced expiratory volume in 1 second, systolic blood pressure, and seven blood biomarkers, with four overlapping biomarkers (albumin, alkaline phosphatase, creatinine, and C-reactive protein) shared with PhenoAge. Additional biomarkers of KDM-BA included blood urea nitrogen, glycated hemoglobin, and total cholesterol, integrated with chronological age (Supplementary File).

Biological age acceleration (BioAgeAccel) is defined as residual between estimated biological aging and chronological age. This metric quantifies biological age deviation adjusted for chronological age and reflects the discordance between an individual’s physiological state and their chronological age, with a positive value of BioAgeAccel indicating accelerated aging (older), whereas a negative value suggesting decelerated aging (younger). PhenoAge acceleration (PhenoAgeAccel) and KDM-BA acceleration (KDM-BA-Accel) were the targeted predictors in the primary analyses.

### Covariates selection

Covariates included chronological age, sex, ethnicity, body mass index (BMI), educational attainment, smoking status, alcohol consumption, income, physical activity (PA), sleep mode (Zou, et al., 2025), history of hypertension, and pharmacological treatments (primarily antihypertensive, lipid-lowering, and glucose-lowering medications), and were systematically assessed for their potential influence on the onset and progression of CKMDs and dementia (Table S1). Considering the highly genetic predisposition to dementia, we also incorporated the polygenic risk score (PRS) for dementia (Supplementary File). Missing covariate data were addressed using Multiple Imputation by Chained Equations method (van Buuren and Groothuis-Oudshoorn, 2011).

### Statistical analyses

#### Multistate analysis of BioAgeAccel and disease progression trajectories (transition pattern A)

The multistate model with Markov proportional hazards was used to examine BioAgeAccel in relation to disease progression from health to CKMDs, then to CKMM or dementia, and finally to death (Putter, et al., 2007). Firstly, based on the limited sample size at the first follow-up of the UK Biobank cohort (2012∼2013; *N*=∼20000), we observed moderate correlations between biological aging measures assessed at baseline and those at the first follow-up (Figure S1), indicating that biological age exhibits good temporal stability. According to the natural course of disease progression, we established a disease trajectory pattern comprising five distinct states with nine transition processes (transition pattern A, Figure 2): (I) healthy to the first occurrence of CKMD (FCKMD); (II) healthy to dementia; (III) healthy to death; (IV) FCKMD to CKMM; (V) FCKMD to dementia; (VI) FCKMD to death; (VII) CKMM to dementia; (VIII) CKMM to death; (IX) dementia to death. We only considered the time of first entry into each state and did not allow transitions back to previous states. Following a prior study (Pan, et al., 2025), for individuals diagnosed with two diseases or death on the same date (PhenoAge: *N*=4,852; KDM-BA: *N*=4,031), we operationally defined the theoretical entry date for the preceding state as 0.5 days before the subsequent state’s date. Models were adjusted for the aforementioned covariates, and hazard ratios (HRs) with 95% confidence intervals (CIs) were reported accordingly.

**Figure 2.**
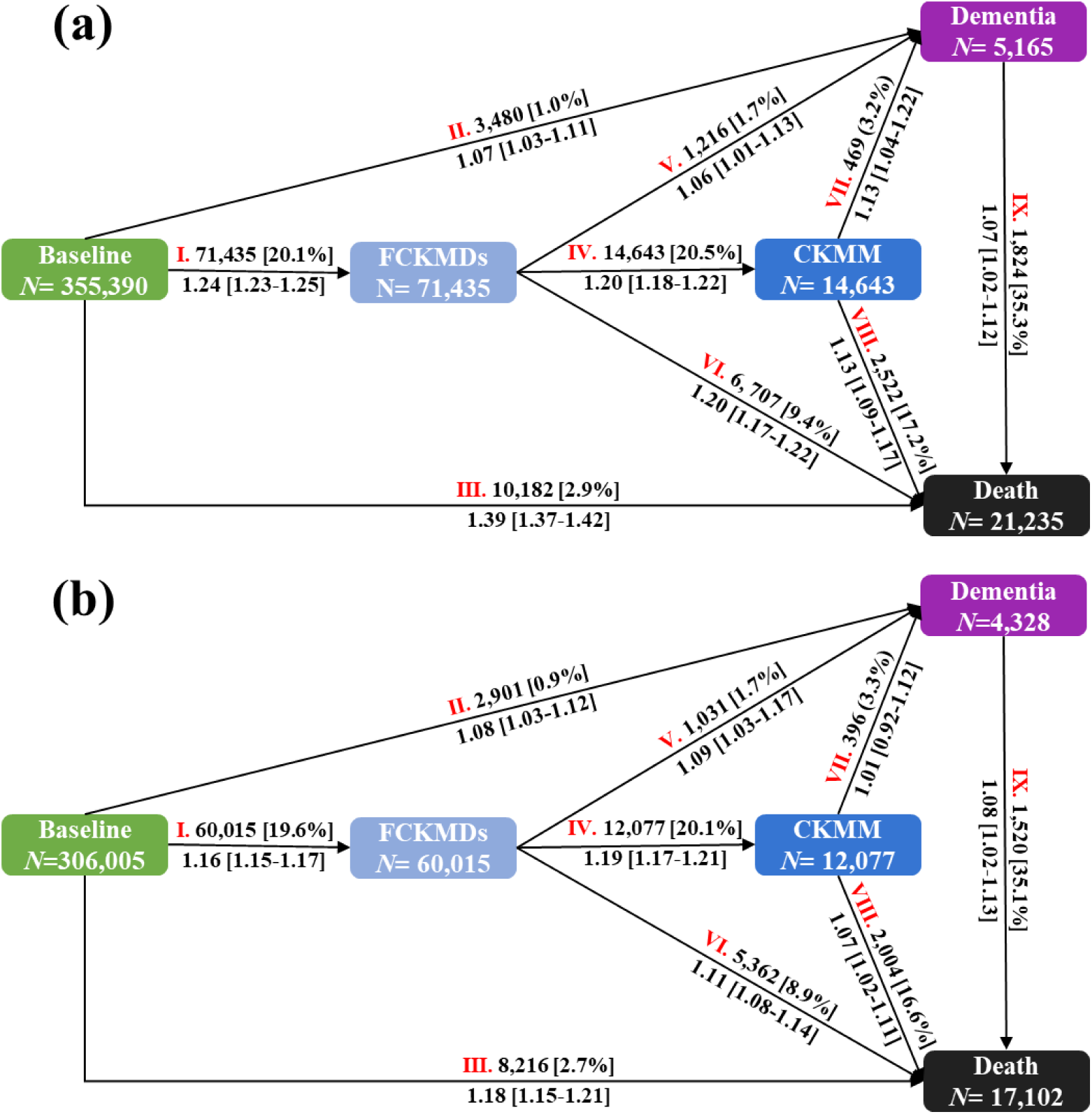
(a) Schematic representation of the multistate models for transition pattern A by PhenoAgeAccel. (b) Schematic representation of the multistate models for transition pattern A by KDM-BA-Accel. Transitions from healthy to FCKMD, CKMM, dementia, and mortality; state-specific number of events was reported in boxes, and the transition-specific number of events and percentages (within brackets) were reported on arrows. The hazard ratios for each transition, along with their 95% confidence intervals, are displayed below the arrows. FCKMDs, the first occurrence of cardiovascular-kidney-metabolic disease; CKMM, cardiovascular-kidney-metabolic multimorbidity. Models were adjusted for chronological age, gender, ethnicity, BMI, educational attainment, smoking status, alcohol consumption, income, PA, sleep mode, history of hypertension, pharmacological treatments, and PRS for dementia.

#### Cumulative transition probabilities, disease progression time and life expectancy by BioAgeAccel quartiles (transition pattern A)

Participants were stratified into quartiles (Q1-Q4) according to their levels of BioAgeAccel, and cumulative transition probabilities across different health states were assessed for each group. Based on the longest follow-up duration, the restricted mean survival time (RMST) was used to estimate the average transition time between states (Uno, et al., 2014). Transitions from healthy to diseases or between disease states were quantified as disease progression time, while transitions to death were defined as life expectancy. Using the lowest BioAgeAccel quartile (Q1) as the reference, differences in disease progression time and life expectancy were compared across higher quartiles to evaluate the potential loss in healthy and overall lifespan attributable to accelerated aging. RMST, defined as the area under the survival curve up to a specified time point, provides greater stability and interpretability than median or mean survival time (Han and Jung, 2022).

#### Association between BioAgeAccel and disease progression trajectories (transition pattern B)

To determine which types of CKMDs were more strongly associated with dementia progression, we further categorized CKMDs into four separate diseases (T2D, CKD, CVD, and stroke) based on the framework of transition pattern A. Thus, in addition to transition processes II, III, VII, VIII, and IX, processes I, IV, V, and VI from pattern A were further subdivided into 12 transition processes, resulting in a disease trajectory model with 8 distinct states and 21 transition processes (transition pattern B, Figure S2-S3). Since the temporal sequence of disease onset could not be determined, for participants diagnosed with at least two CKMDs on the same date, we assumed the order of occurrence to be T2D, followed by CKD, CVD, and stroke (Jiang, et al., 2024; Ndumele, et al., 2023). To account for this, we subtracted 0.5 days from the recorded date of the preceding disease.

#### Stratified analyses and sensitivity analyses

To explore potential effect modifiers, we further stratified the multistate analyses of transition pattern A based on gender, chronological age (>65 years or ≤60 years), educational attainment (college degree vs. non-college degree), income (<£31,000 vs. ≥£31,000), smoking status (never vs. former or current smoker), alcohol consumption (more than once per week vs. less than once per week), PA (low-to-moderate vs. high), or sleep patterns (healthy vs. unhealthy).

Additionally, four sensitivity analyses were conducted to validate the robustness of our findings with respect to biological aging measures: (1) For participants who transitioned between different states on the same day, to assess the impact of time interval selection on the results, we applied time intervals of 0.5, 1, and 3 years to estimate the entry date of their previous state; (2) Participants who experienced multiple state transitions within the same day were excluded; (3) Events occurring within 2 years of follow-up initiation were excluded to reduce potential reverse causality; and (4) To examine the possible impact of racial differences on the results, the multistate model for transition pattern A was reconstructed using data from white participants only.

#### Statistical software

All statistical analyses were performed under the R software (version 4.2.3). All statistical tests were two-sided, and *P*<0.05 were considered as statistically significant. Multiple-imputation was achieved by the mice package (van Buuren and Groothuis-Oudshoorn, 2011). The calculation of biological aging was implemented through the BioAge package (Kwon and Belsky, 2021). The Irwin’s RMST was performed by the survRM2 package (Tian, et al., 2014; Uno, et al., 2014). The multistate models were performed by the mstate package (Putter, et al., 2007).

## Results

### Baseline characteristics

There were 355,390 participants retained for PhenoAge-relevant analyses and 306,005 participants kept for KDM-BA-relevant analyses (Figure 1). The average age of these participants was 56±8 years, 57% were female, and the majority of them (>94%) were White (Table S2). Individuals transition to FCKMD, CKMM, dementia or death during follow-up (median follow-up of 13.4 [interquartile range 12.7-14.1] years) exhibited differences in baseline characteristics compared to those without disease progression, including older age, a higher proportion of males, higher BMI, greater exposure to tobacco, lower income levels, reduced PA, poorer sleep health, and a higher likelihood of having a history of hypertension and using cardiometabolic medications.

### Association between BioAgeAccel and disease progression trajectories in transition pattern A

Figure 2 illustrates the progression across diseases in transition pattern A, as captured by two biological aging measures. The multistate analysis revealed that both BioAgeAccel indicators were significantly positively associated with nearly all stages of disease progression except for the transition from CKMM to dementia, where KDM-BA-Accel did not show a statistically significant link (*P*=0.77). First, in the dynamic progression of CKMM, the risk of PhenoAgeAccel in the transition from healthy to FCKMD (I) was approximately 8% greater compared to KDM-BA-Accel; while the risk in the transition from FCKMD to CKMM (IV) was nearly consistent for them (HR=1.20 [1.18-1.22] vs. 1.19 [1.17-1.21]).

Second, in the dementia-related transitions (□, □, □), the risk of PhenoAgeAccel in the path from CKMM to dementia (HR=1.13 [1.04-1.22]) was slightly higher than that in the transition from healthy to dementia (HR=1.07 [1.03-1.11]) or from FCKMD to dementia (HR=1.06 [1.01-1.13]); but opposite phenomena were observed for KDM-BA-Accel. Third, in the four transition paths (□, □, □, □) toward mortality, PhenoAgeAccel typically exhibited stronger risk effects than KDM-BA-Accel. Specifically, PhenoAgeAccel was associated with increased mortality risk in the paths from healthy (HR=1.39 [1.37-1.42]), FCKMD (HR=1.20 [1.17-1.22]), or CKMM (HR=1.13 [1.09-1.17]), with an approximately 21%, 9%, and 6% higher risk than the corresponding impacts of KDM-BA-Accel, respectively. Further, the associations between BioAgeAccel and multiple disease transitions exhibited dose-response patterns (Figure 3a and Table S3).

**Figure 3.**
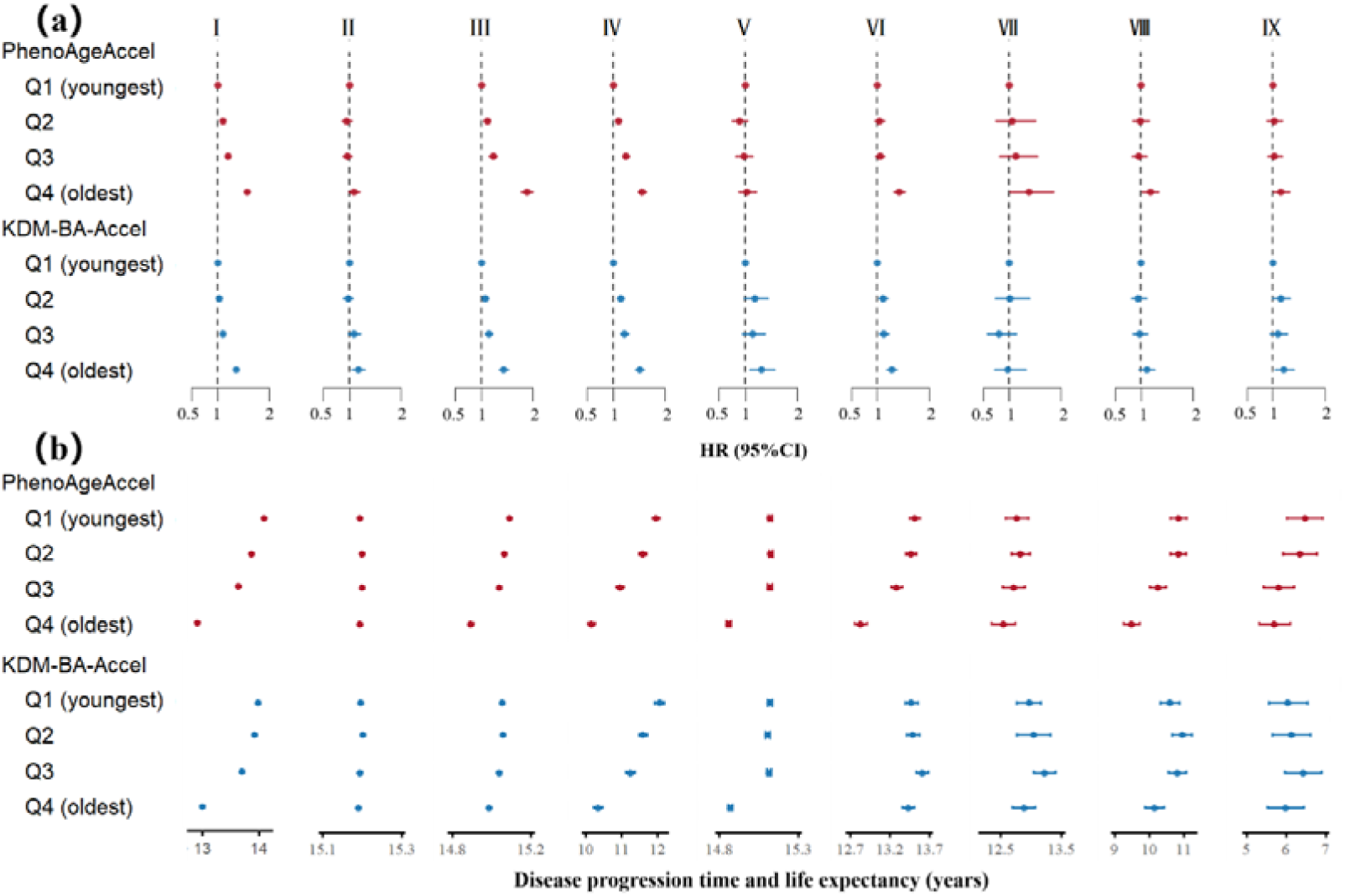
(a) Associations between BioAgeAccel (quartiles) and the transitions from healthy to FCKMDs, CKMM, dementia, and mortality. (b) Disease progression time and life expectancy among disease transitions with different levels of BioAgeAccel (quartiles). PhenoAgeAccel, PhenoAge acceleration; KDM-BA-Accel, KDM-BA acceleration; Q1-Q4, Quartile 1 (the first quartile)-Quartile 4 (the fourth quartile).

### Impact of BioAgeAccel on cumulative transition probabilities, disease progression time and life expectancy in transition pattern A

We observed significant differences in cumulative transition probabilities for progression to FCKMD, CKMM, and mortality across different BioAgeAccel levels (Figures S4-S5). Compared with individuals in the lowest quartile (Q1) of BioAgeAccel, those in higher quartiles (Q2-Q4) demonstrated significantly increased cumulative transition probabilities (all *P*<0.05). However, no significant differences in the cumulative transition probabilities to dementia were observed across different BioAgeAccel levels.

Further RMST analysis discovered that, compared to individuals in the lowest quartile of BioAgeAccel (Q1), those in higher quartiles (Q2-Q4) exhibited significantly shorter disease progression time and life expectancy at certain stages of disease progression (Figure 3b and Table S4). For instance, individuals in the highest quartile (Q4) experienced a reduction in disease progression time from healthy to FCKMD of 1.18 [1.15-1.22] years for PhenoAge and 0.99 [0.95-1.02] years for KDM-BA. The transition from FCKMD to CKMM was shortened by 1.79 [1.64-1.94] years for PhenoAge and 1.71 [1.53-1.89] years for KDM-BA. Regarding life expectancy, individuals with CKMDs in Q4 had shorter life spans by 0.68 [0.58-0.79] years for PhenoAge compared to Q1. Among those with CKMM, the reductions were 1.36 [1.03-1.69] years for PhenoAge and 0.44 [0.06-0.83] years for KDM-BA, while among those with dementia, the decreases were 0.77 [0.16-1.38] years for PhenoAge. Additionally, although slight reductions in disease progression time toward dementia were observed, they did not reach statistical significance (all *P*>0.05), consistent with the relatively weaker effects.

### Association between BioAgeAccel and disease progression trajectories (transition pattern B)

During the transition from healthy to FCKMD and subsequently to CKMM, both PhenoAgeAccel and KDM-BA-Accel were significantly related to increased progression risk (Table 1). Individuals with CVD as the initial diagnosis exhibited a higher risk of progressing to CKMM (PhenoAge: HR=1.25 [1.22-1.28]; KDM-BA: HR=1.23 [1.19-1.26]) compared to those initially diagnosed with CKD, T2D, or stroke.

**Table 1.**
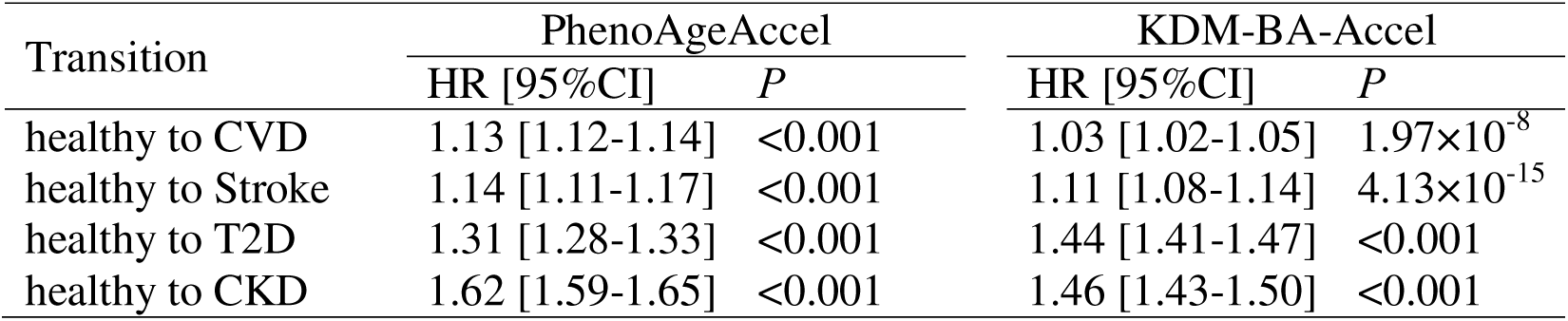
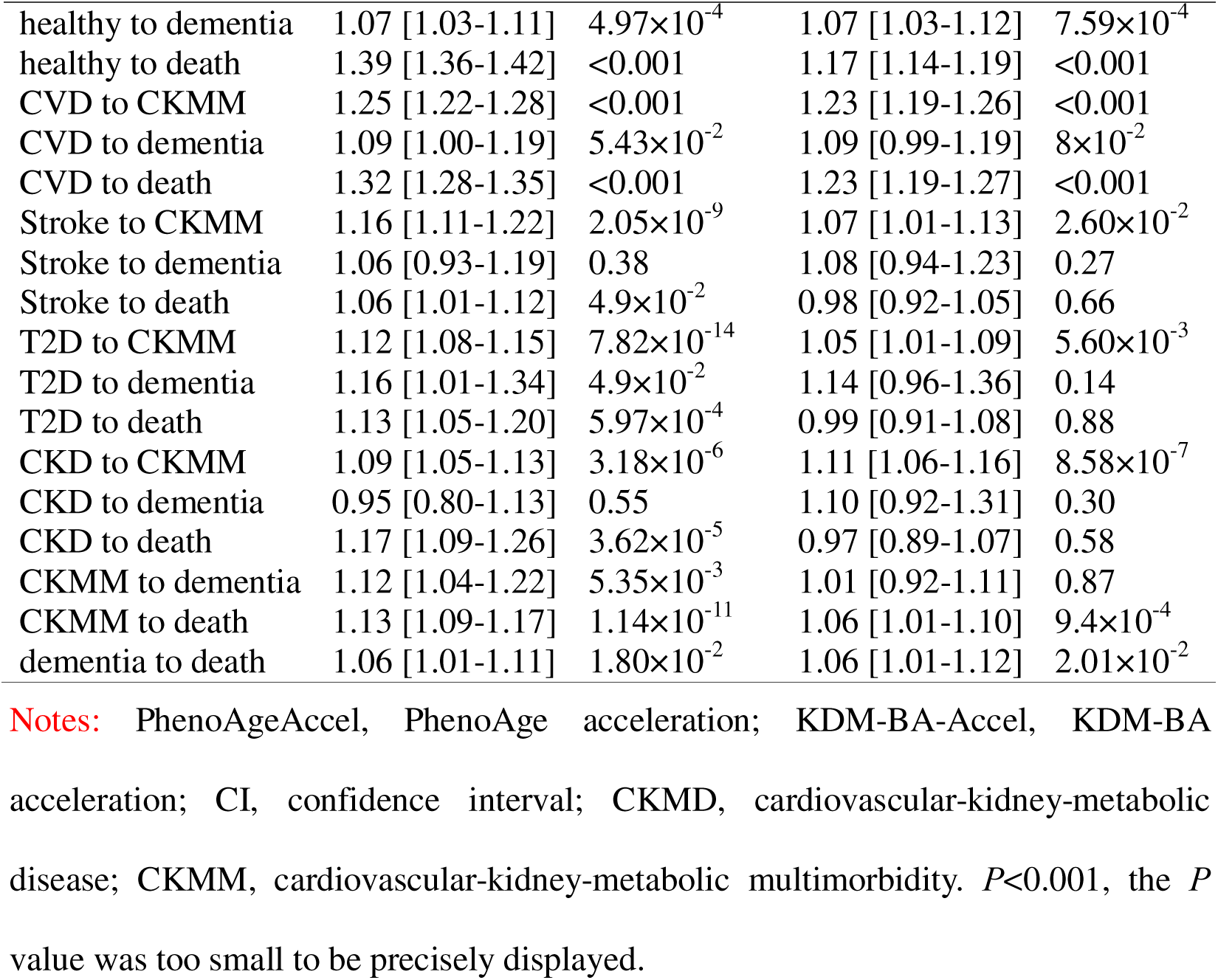
Associations of BioAgeAccel with different transitions from baseline to specific CKMD, CKMM, dementia and death.

| Transition | PhenoAgeAccel |  | KDM-BA-Accel |  |
| --- | --- | --- | --- | --- |
|  | HR [95%CI] | *P* | HR [95%CI] | *P* |
| healthy to CVD | 1.13 [1.12-1.14] | <0.001 | 1.03 [1.02-1.05] | $1.97 \times 10^{-8}$ |
| healthy to Stroke | 1.14 [1.11-1.17] | <0.001 | 1.11 [1.08-1.14] | $4.13 \times 10^{-15}$ |
| healthy to T2D | 1.31 [1.28-1.33] | <0.001 | 1.44 [1.41-1.47] | <0.001 |
| healthy to CKD | 1.62 [1.59-1.65] | <0.001 | 1.46 [1.43-1.50] | <0.001 |
| healthy to dementia | 1.07 [1.03-1.11] | $4.97 \times 10^{-4}$ | 1.07 [1.03-1.12] | $7.59 \times 10^{-4}$ |
| healthy to death | 1.39 [1.36-1.42] | <0.001 | 1.17 [1.14-1.19] | <0.001 |
| CVD to CKMM | 1.25 [1.22-1.28] | <0.001 | 1.23 [1.19-1.26] | <0.001 |
| CVD to dementia | 1.09 [1.00-1.19] | $5.43 \times 10^{-2}$ | 1.09 [0.99-1.19] | $8 \times 10^{-2}$ |
| CVD to death | 1.32 [1.28-1.35] | <0.001 | 1.23 [1.19-1.27] | <0.001 |
| Stroke to CKMM | 1.16 [1.11-1.22] | $2.05 \times 10^{-9}$ | 1.07 [1.01-1.13] | $2.60 \times 10^{-2}$ |
| Stroke to dementia | 1.06 [0.93-1.19] | 0.38 | 1.08 [0.94-1.23] | 0.27 |
| Stroke to death | 1.06 [1.01-1.12] | $4.9 \times 10^{-2}$ | 0.98 [0.92-1.05] | 0.66 |
| T2D to CKMM | 1.12 [1.08-1.15] | $7.82 \times 10^{-14}$ | 1.05 [1.01-1.09] | $5.60 \times 10^{-3}$ |
| T2D to dementia | 1.16 [1.01-1.34] | $4.9 \times 10^{-2}$ | 1.14 [0.96-1.36] | 0.14 |
| T2D to death | 1.13 [1.05-1.20] | $5.97 \times 10^{-4}$ | 0.99 [0.91-1.08] | 0.88 |
| CKD to CKMM | 1.09 [1.05-1.13] | $3.18 \times 10^{-6}$ | 1.11 [1.06-1.16] | $8.58 \times 10^{-7}$ |
| CKD to dementia | 0.95 [0.80-1.13] | 0.55 | 1.10 [0.92-1.31] | 0.30 |
| CKD to death | 1.17 [1.09-1.26] | $3.62 \times 10^{-5}$ | 0.97 [0.89-1.07] | 0.58 |
| CKMM to dementia | 1.12 [1.04-1.22] | $5.35 \times 10^{-3}$ | 1.01 [0.92-1.11] | 0.87 |
| CKMM to death | 1.13 [1.09-1.17] | $1.14 \times 10^{-11}$ | 1.06 [1.01-1.10] | $9.4 \times 10^{-4}$ |
| dementia to death | 1.06 [1.01-1.11] | $1.80 \times 10^{-2}$ | 1.06 [1.01-1.12] | $2.01 \times 10^{-2}$ |

**Notes:** PhenoAgeAccel, PhenoAge acceleration; KDM-BA-Accel, KDM-BA acceleration; CI, confidence interval; CKMD, cardiovascular-kidney-metabolic disease; CKMM, cardiovascular-kidney-metabolic multimorbidity. $P < 0.001$ , the $P$ value was too small to be precisely displayed.

Regarding dementia-related transitions, PhenoAgeAccel showed significant associations across three pathways: from healthy (HR=1.07 [1.03-1.11]), T2D (HR=1.16 [1.01-1.34]), and CKMM (HR=1.12 [1.04-1.22]). In contrast, KDM-BA-Accel was only significantly associated with the transition from healthy (HR=1.07 [1.03-1.12]) to dementia. Additionally, individuals with T2D (PhenoAge: HR=1.16 [1.00-1.34]; KDM-BA: HR=1.14 [0.96-1.36]) had a higher likelihood of progressing to dementia than those with CKD, CVD, or stroke (Table 1).

In terms of mortality-related transitions, PhenoAgeAccel was significantly linked to enhanced mortality risks across all pathways, whereas KDM-BA-Accel showed no significant associations in several mortality transitions, such as from stroke (*P*=0.66), T2D (*P*=0.88), and CKD (*P*=0.58) to death (Table 1).

### Results of stratified analyses

Subgroup analyses revealed heterogeneity in the effects of BioAgeAccel on disease progression (Tables S5-S6). The impact of BioAgeAccel on dementia progression was more pronounced among younger individuals (≤60 years). Specifically, PhenoAgeAccel significantly increased the risk of transitioning to dementia from healthy (HR=1.20 [1.10-1.30]), FCKMD (HR=1.28 [1.10-1.48]), and CKMM (HR=1.26 [1.02-1.55]) in younger group. In contrast, these associations were markedly attenuated in the older group (>60 years), with smaller and insignificant risk estimates (all *P*>0.05). For mortality-related transitions, PhenoAgeAccel was related to approximately 11%, 12%, and 10% higher risks of death from the healthy, CKMM, and dementia, respectively, in the younger group compared to the older group. Similar patterns were observed for KDM-BA.

PA showed state-specific moderating effects. Among individuals engaging in high-intensity PA, BioAgeAccel showed much weaker and insignificant associations with dementia progression from healthy (KDM-BA: HR=1.02 [0.96-1.09]) and from FCKMD (PhenoAge: HR=1.02 [0.92-1.13]; KDM-BA: HR=1.04 [0.94-1.15]). Conversely, those with low-to-moderate PA showed higher risks (healthy-to-dementia: KDM-BA HR=1.12 [1.06-1.18]; FCKMD-to-dementia: PhenoAge HR=1.09 [1.01-1.17]; KDM-BA HR=1.13 [1.04-1.23]).

Additionally, higher educational attainment and healthier lifestyles (including non-smoking, low alcohol consumption, and healthy sleep) significantly mitigated the effects of BioAgeAccel on transitions to FCKMD, dementia, and mortality. However, in the CKMM-to-dementia transition, individuals with healthier lifestyles (non-smoking, low alcohol consumption) exhibited a stronger association between BioAgeAccel and increased dementia risk.

### Results of sensitivity analyses

The associations between BioAgeAccel and disease progression risk remained largely stable when assigning different time intervals for the same entry date (Tables S7-S9). After excluding individuals with multiple state transitions occurring on the same day (HR=1.06 [1.00-1.13], *P*=0.062) or those who progressed to FCKMD, CKMM, dementia, or death within the first two years of follow-up (HR=1.06 [1.00-1.13], *P*=0.068), the association between PhenoAgeAccel and the transition from FCKMD to dementia was attenuated to a borderline significance level (Tables S10-S11). Additionally, the associations between BioAgeAccel and disease progression remained consistent among White participants (Table S12).

## Discussion

### Summary of our Study

This study systematically evaluated the association between BioAgeAccel and disease transitions, ranging from healthy to FCKMD, then progression to CKMM, dementia, and ultimately to death using prospective cohort data from the UK Biobank. We revealed that BioAgeAccel was significantly associated with all disease transitions, reducing transition times to subsequent disease states to different extents. The robustness of these findings was supported by long-term follow-up, large samples, two complementary BioAgeAccel measures and extensive sensitivity analyses.

### Comparison with previous studies

#### BioAgeAccel and disease progression: novel insights from multistate modeling

Although the association between BioAgeAccel and the risk of single CKMD and dementia was widely documented (Costantino, et al., 2016; Ye, et al., 2024), its role in disease progression remains insufficiently explored. A recent study indicated PhenoAgeAccel was closely associated with both the onset of T2D and the progression of its complications (Pan, et al., 2025); others demonstrated BioAgeAccel significantly increased risks of initial cardiometabolic diseases (CMD) and subsequent multimorbidity (Jiang, et al., 2024). Additionally, He et al. examined the link between BioAgeAccel and the development of CMD and progression to dementia (He, et al., 2024). However, the use of separate Cox proportional hazard models to evaluate each outcome path in those studies, without incorporating a unified dynamic framework or accounting for competing risks, limits the applicability of their findings.

In contrary to existing research, our study employed a multistate model, focusing on two major chronic conditions — CKMDs and dementia — to construct the first dynamic transition framework from CKMD onset to multimorbidity, dementia, and death, and assessed the role of BioAgeAccel in these progression processes. Consistent with previous work (Wang, et al., 2025; Zheng, et al., 2025), we found that BioAgeAccel was significantly linked with increased risk of FCKMD and accelerated its onset by an average of 1.09 years. BioAgeAccel was also related to higher risk of FCKMD progressing to CKMM, with an average advancement of 1.75 years in disease onset. These findings suggest that BioAgeAccel serves not only as a predictor of disease onset but also plays a critical role in the formation of multimorbidity. Regarding health outcomes, individuals with single CKMD and CKMM exhibited substantially greater mortality risk compared to the healthy population (2.7∼2.9%), underscoring the value of identifying high-risk individuals using BioAgeAccel. Implementing targeted primary and secondary prevention strategies in these populations may help delay or prevent disease progression, reduce the risk of multimorbidity, and ultimately lower mortality rates.

Regarding dementia progression, building upon prior work (Liu, et al., 2023; Sugden, et al., 2022), we further demonstrated that BioAgeAccel significantly facilitated the transition from FCKMD/CKMM to dementia, supporting its potential role as a shared driver of CKMDs and neurodegenerative diseases. Notably, the impact of PhenoAgeAccel on the transition from CKMM to dementia was greater than that from healthy state to dementia, highlighting the need for special attention to individuals with high BioAgeAccel within the CKMD population, particularly among those with CKMM. Tailored cognitive decline intervention strategies for these high-risk groups may be warranted.

#### Insights from transition pattern B and key monitoring populations

Results from transition pattern B further indicated that individuals with BioAgeAccel combined with CVD or T2D represented key monitoring populations for preventing CKMM or dementia, respectively. We demonstrated that patients with first-onset CVD exhibited a higher risk of progressing to CKMM compared to those with first-onset CKD, T2D, or stroke. As a systemic disorder, CVD typically involves activation of pathological states such as atherosclerosis, systemic inflammation, and vascular endothelial dysfunction (Frostegård, 2013), which not only impair the cardiovascular system but also exacerbate renal function decline (Ronco, et al., 2008) and glucose metabolism abnormalities (Li, et al., 2022) through hemodynamic disturbances and oxidative stress, ultimately accelerating multisystem deterioration. Additionally, CVD patients frequently present with comorbidity burdens (e.g., hypertension, hyperuricemia, low high-density lipoprotein cholesterol) (Petrie, et al., 2018; Zhang, et al., 2019), which signify elevated pathological loading during early CVD stages, thereby markedly accelerating progression toward CKMM multimorbidity. BioAgeAccel, closely linked to atherosclerosis and inflammation, further amplifies these adverse effects (Kennedy, et al., 2014; Wang and Bennett, 2012). Although CKD, T2D, and stroke were also related to biological aging, their systemic impacts were comparatively weaker, making CVD a stronger catalyst for multimorbidity under BioAgeAccel.

Further, we found that FCKMD patients with T2D as the initial diagnosis showed a higher likelihood of progressing to dementia than those with CKD, CVD, or stroke. The hyperglycemic state and insulin resistance induced by T2D impair the blood-brain barrier, disrupt insulin signaling, and activate neuroinflammation, promoting neurodegeneration (Banks and Rhea, 2021). Concurrently, BioAgeAccel aggravates inflammation and immune senescence, weakening cognitive resilience and further intensifying T2D-related dementia progression (Shadyab, et al., 2022). Additionally, PhenoAgeAccel exhibits superior predictive capacity for mortality progression compared to KDM-BA-Accel, potentially attributable to the inherent design of PhenoAge metrics to specifically reflect and predict mortality risks (Liu, et al., 2018).

#### Subgroup analyses: moderating factors and intervention windows

Largely consistent with a prior finding (Ye, et al., 2024), our work demonstrated a marked age-specific effect of BioAgeAccel on disease progression: its impact on dementia development was more pronounced among middle-aged individuals (≤60 years) than in older adults (>60 years), suggesting that midlife may represent a critical window for implementing anti-aging interventions aimed at slowing dementia progression. Moreover, smoking and alcohol consumption exacerbated the detrimental effects of BioAgeAccel, in line with recent evidence (Zhang, et al., 2024). Beyond lifestyle factors, educational attainment also emerged as a significant buffer; individuals with university degree exhibited lower risk of progressing from CKMD to dementia, possibly due to greater cognitive reserve (Yang, et al., 2024).

Importantly, while prior studies have consistently highlighted the protective role of PA in preventing dementia (Ning, et al., 2025; Vidyanti, et al., 2025), our study further extends this understanding by revealing that high-intensity PA in healthy individuals or those with a single CKMD condition may significantly attenuate the adverse association between BioAgeAccel and dementia. However, this protective effect of high-intensity PA was no longer significant in individuals with CKMM. Intriguingly, in the CKMM-to-dementia transition, we observed a paradoxical phenomenon where BioAgeAccel exhibited more pronounced adverse effects among non-smokers and individuals with low alcohol consumption. This counterintuitive association likely indicates the potential involvement of yet-unclear biological mechanisms or compensatory threshold effects, warranting rigorous investigation in future studies to elucidate the underlying pathophysiological processes.

### Public health implications of our findings

This study reveals the complex relations between CKMDs, dementia, and biological aging, providing strong theoretical support for dynamic monitoring and prevention against these diseases and conditions, as well as the formulation of public health policies. First, our study supports the use of BioAgeAccel as a tool for monitoring healthy aging and risk stratification interventions (Jiang, et al., 2024). By assessing BioAgeAccel, we can effectively identify individuals at higher risk of disease progression, particularly those who have already exhibited early signs of aging. This allows for more precise risk prediction and the optimization of health management and intervention strategies.

Second, middle-aged individuals should be prioritized for aging monitoring (Livingston, et al., 2024). Early intervention with BioAgeAccel can effectively slow down or even reverse the aging process. Implementing biological age assessment and early intervention measures in middle-aged populations represents a potentially effective pathway to reduce the incidence rate of dementia. Finally, the role of a healthy lifestyle in disease prevention is crucial. In addition to promoting basic public health knowledge such as smoking cessation and reduced alcohol consumption, communities and hospitals should encourage healthy individuals and those with a single CKMD to engage in high-intensity physical activity. In regions with lower educational attainment, policies to promote higher education should be strengthened to enhance individuals’ resilience against cognitive decline.

### Strengths and limitations of this study

This study has several notable strengths. First, to our knowledge, it represents the first systematic application of multistate models (Putter, et al., 2007) to evaluate the associations between biological aging and the dynamic transitions from FCKMD onset to subsequent states of CKMM, dementia, and mortality. It is also the first to quantify the impact of BioAgeAccel on the disease progression time at each stage, providing a comprehensive assessment of the role of BioAgeAccel in disease progression. Methodologically, compared to traditional survival analysis approaches, multistate models not only allow examination of multiple intermediate health states and their transition pathways but also account for competing risks, thereby more accurately reflecting the complexity and dynamism of disease progression. Second, the concurrent use of two distinct biological age algorithms — PhenoAge (Liu, et al., 2018) and KDM-BA (Klemera and Doubal, 2006) — enabled consistency validation and cross-comparison of results, significantly enhancing the robustness and credibility of our conclusions. Third, leveraging the large-scale, prospective design of the UK Biobank cohort not only provides sufficient statistical power but also allows for better clarification of temporal relationships between exposures and outcomes (Sudlow, et al., 2015), thereby reducing the potential for reverse causation bias. Finally, we conducted a series of sensitivity analyses to assess the robustness of our findings, and incorporated a wide range of covariates to minimize confounding and explore potential effect modification.

Nevertheless, several limitations should be acknowledged. First, the absence of repeated measurements precluded the assessment of longitudinal changes in biological aging, thereby limiting deeper understanding of how dynamic aging trajectories influences disease progression. Second, for participants reporting multiple events on the same day, we relied on empirical assumptions to determine event ordering. This may reduce the strength of multistate models, but only very slightly, since the number of cases with this condition was relatively small (about 6.8% of FCKMD cases). Third, various aging indicators generally reflect distinct physiological and biochemical states (López-Otín, et al., 2023; Rutledge, et al., 2022); validations are thus warranted for biological aging measures beyond PhenoAge and KDM-BA. Other limitations include the healthy volunteer bias inherent in the UK Biobank cohort (Fry, et al., 2017), as well as the predominance of middle-aged and older White individuals in the sample, which may introduce bias in risk estimation and limit the generalizability of our findings to other racial and age groups (Sudlow, et al., 2015).

## Conclusions

Based on the large-scale prospective UK Biobank cohort, this study revealed the significant promotive role of BioAgeAccel in the onset of CKMD and their subsequent progression to CKMM, dementia, and mortality. These findings underscore the potential utility of BioAgeAccel as a tool for monitoring healthy aging and informing risk stratification strategies, offering a means to identify high-risk individuals exhibiting early signs of accelerated aging and thereby enabling accurate risk prediction. Our findings also suggest that implementing biological age assessments in middle-aged populations, in conjunction with the promotion of healthy lifestyle interventions, represents a promising strategy for effectively mitigating the burden of CKMDs and dementia.

## Declarations

### Ethics approval and consent to participate

The UK Biobank had approval from the North West Multi-Centre Research Ethics Committee (MREC) as a Research Tissue Bank (RTB) approval. All participants provided written informed consent before enrolment in the study, which was conducted in accordance with the Declaration of Helsinki. This approval means that researchers do not require separate ethical clearance and can operate under the RTB approval.

## Consent for publication

All authors have approved the manuscript and given their consent for submission and publication.

## Data availability

All data generated or analyzed during this study are included in this published article and its supplementary information files. This study used the UK Biobank resource with the application ID 88159. Researchers can access to the UK Biobank by submitting an application to the UK Biobank official website (https://www.ukbiobank.ac.uk/).

## Disclosure

The authors declare that the research was conducted in the absence of any commercial or financial relationships that could be construed as a potential conflict of interest.

## Author’s contributions

PZ conceived the idea for the study. PZ obtained the data. GJY cleared up the datasets; GJY performed the data analyses. PZ and GJY interpreted the results of the data analyses. PZ and GJY wrote the manuscript.

## Abbreviations

CKMDs: Cardiovascular-kidney-metabolic diseases
T2D: Type 2 diabetes
CKD: Chronic kidney disease
CVD: Cardiovascular disease
CKMM: Cardiovascular-kidney-metabolic multimorbidity
KDM-BA: Klemera-Doubal Method Biological Age
BioAgeAccel: Biological age acceleration
PhenoAgeAccel: PhenoAge acceleration
KDM-BA-Accel: Klemera-Doubal Method Biological Age acceleration
BMI: Body mass index
PA: Physical activity
PRS: Polygenic risk score
FCKMD: First occurrence of Cardiovascular-kidney-metabolic disease
HRs: Hazard ratios
CIs: confidence intervals
Q1: quartiles 1
Q2: quartiles 2
Q3: quartiles 3
Q4: quartiles 4
RMST: Restricted mean survival time
CMD: Cardiometabolic diseases

## Acknowledgements

This study was mainly based on the UK Biobank resource under application number 88159. The UK Biobank was established by the Wellcome Trust medical charity, Medical Research Council, Department of Health, Scottish Government, and the Northwest Regional Development Agency. It has also had funding from the Welsh Assembly Government, British Heart Foundation and Diabetes UK. The data analyses in the present study were carried out with the high-performance computing cluster that was supported by the special central finance project of local universities for Xuzhou Medical University.

